# Is There a Correlation Between Pulmonary Inflammation Index With COVID-19 Disease Severity And Outcome?

**DOI:** 10.1101/2020.09.09.20182592

**Authors:** Aliae AR Mohamed Hussein, Islam Galal, Mohammed M Mohammed, Howida K Abd Elaal, Karim Aly

## Abstract

**Rational:** the radiologic pulmonary inflammatory index (PII) may be used as early predictor of inflammation as laboratory assessments in COVID-19 cases. **The purpose of this study was to** compare the clinical and radiological features between the cases of COVID-19 necessitating admittance to the intensive care unit (ICU) and those who did not, and to correlate the radiological pulmonary inflammation index (PII) with other inflammatory markers and outcome.

**Patients and methods:** This study included 72 patients consecutively admitted with confirmed COVID-19. Their electronic records of were retrospectively revised and the demographic, clinical, laboratory (complete blood count, C reactive protein, D dimer and serum ferritin), HRCT data, pulmonary inflammation index (PII) and the outcomes of the patients (ICU admission, death, recovery, and referral) were analyzed.

**Results:** They were 50/50% males/females, mean age was 47.1 ± 16.8 (median 47 years). During their stay, 15.3% necessitated ICU admittance, 49 (68%) cured and discharged, 9 cases referred and five cases (6.9%) died. The baseline lesions identified were ground glass opacification recognized in (93%), higher PII and >3 lobes affection were considerably recorded in those who required ICU admittance (P= 0.041 and 0.013). There were moderate positive correlations between PII with age (r=0.264, P=0.031) and other prognostic inflammatory indicators as ferritin (r=0.225, P=0.048), D Dimer (r=0.271, P=0.043) and serum creatinine.

**Conclusions:** The use of PII together with clinical and laboratory data may be valuable in defining the inflammatory state of COVID-19. It was correlated with other inflammatory indices as D dimer, ferritin even before clinical deterioration. This may allow clinicians to avoid the progression of the illness and improve cure rates by proper early intervention.

## Introduction

The clinical presentations of COVID-19 range from asymptomatic case to mild, moderate and severe forms rapidly progressing to hospital admission, septicemia or even death in 4-15% of cases (1). So, it is crucial to detect the (COVID-19) disease and its severity as early as possible. Due to the low sensitivity of PCR, several patients with COVID-19 may not be recognized and may not gain proper management in time.

Recently the use of HRCT, as a routine imaging tool for the diagnosis of COVID-19 pneumonia, is comparatively simple to implement a fast diagnosis. As recently reported, chest CT demonstrates typical radiographic features in almost all COVID-19 cases (2) even with cases with preliminary negative PCR results (3,4,5) and the radiologic pulmonary inflammatory index (PII) may be used as early predictor of inflammation as laboratory assessments (6).

***The objective of the current work*** was to compare the clinical and radiological features between the cases of COVID-19 necessitating admittance in the intensive care unit (ICU) and those who did not, and to correlate the radiological pulmonary inflammation index (PII) with the other inflammatory indices and outcome.

### Patients and Methods

This study included all patients consecutively admitted in Aswan university hospital during the period from April to June 2020. All cases had confirmed COVID-19 infection based on PCR, and received standardized treatment protocols of Egyptian Ministry of Health and Population based on disease severity. We excluded children, pregnant females and those with negative PCR results from this study. The electronic records of the patients were retrospectively revised and the demographic data, clinical manifestations, presence of comorbid diseases, laboratory investigations (complete blood count, C reactive protein, D dimer and serum ferritin), and radiological topographies based on HRCT findings, moreover the outcomes of these patients (ICU admission, death, recovery, and referral) were analyzed.

### Chest HRCT

HRCT examination was performed to all the population under study using standard protocol via Aquilion 64, Toshiba 160-slice CT scanner, aquilion TM prime. Non-contrast scans were obtained at full inspiration from the apex to the base of the lung in supine subjects. The acquisition parameters were as follows: Sequential mode, 1-mm collimation and 10-mm interval, 180-260 mA average tube current (depending on body built) and 120-140 kV tube voltage.

All the cuts of CT were appraised by the use of a lung window, with −500 HU window level and 1500 HU window width. The CT images were revised by 2 specialists (pulmonologist) who were blinded to the clinical records of the cases. The CT imaging topographies were entirely evaluated, and the following findings were highlighted: ground glass opacity (GGO), consolidation, crazy paving pattern, interlobular septal thickening, bronchial wall thickening, vascular enlargement, presence of halo sign, reverse halo sign and pleural effusion in harmony with the typical morphologic descriptors built on the Fleischner Society Nomenclature Commission references and comparable studies (**7, 8**). Definitely, the assessment of the size and extent of lung affection was based on the number of segments affected by the disease: normally there are ten subdivisions in the left lung and ten subdivisions in the right lung (two segments were deliberated in the apicoposterior section of the left upper lobe and two segments were deliberated in the inferior front section of the left lower lobe). According to the assessment standard proven by Chongqing Radiologist Association of China, the pulmonary inflammation index (PII) was accomplished for each case using the following formula: [PII = (distribution score + size score)/40*100%]. This score uses lung opacification as a substitute for determination of lungs extension of the disease.

### Statistical analysis

All statistical analyses were performed using the Statistical Package for the Social Sciences (SPSS), version 22.0 (SPSS Inc., Chicago, IL, USA). Descriptive analyses were performed for categorical variables. Continuous variables were expressed as the mean ±SD, median, tested for normality, and compared using the independent samples t-test. Otherwise, the Mann-Whitney test was used. The proportions of categorical variables were compared using a chi-squared test. Pearson product-moment analysis was used to evaluate relations between variables. P < 0.05 was considered statistically significant.

**Ethical consideration:** the study was approved by Ministry of Health and Population and ethical committee Faculty of Medicine, Assiut University, Egypt.

The study is registered in clinicaltrial.gov ID: NCT04479293

## Results

### Clinical data of COVID-19 patients on admission

The current study described the characteristics of COVID-19 cohort of 72 patients admitted in single center. They were 50/50 % males/females, mean age was 47.1 ± 16.8, median (IQR) was 47 years. Most cases (48.6%) were contacts to positive cases. The mean duration of complaints was 4.3 ± 2.8 days before admission. 66.7% of the cases were isolated in wards, 18.1% requested home isolation, while 15.3% necessitated ICU admission. The recorded outcome was: 49 (68%) of 72 patients discharged, 9 cases referred (12, 5%) and five (6.9%) cases died. Regarding the clinical severity classification of the disease (according to WHO), 50% of cases were moderate in severity. The associated comorbidities, clinical symptoms and laboratory data were recorded in ***Table 1***.

**Table 1.**
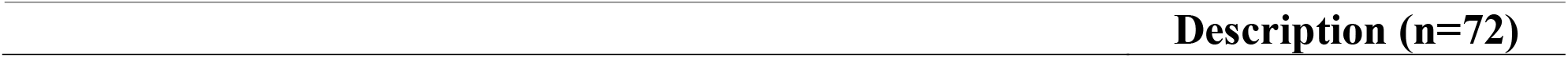
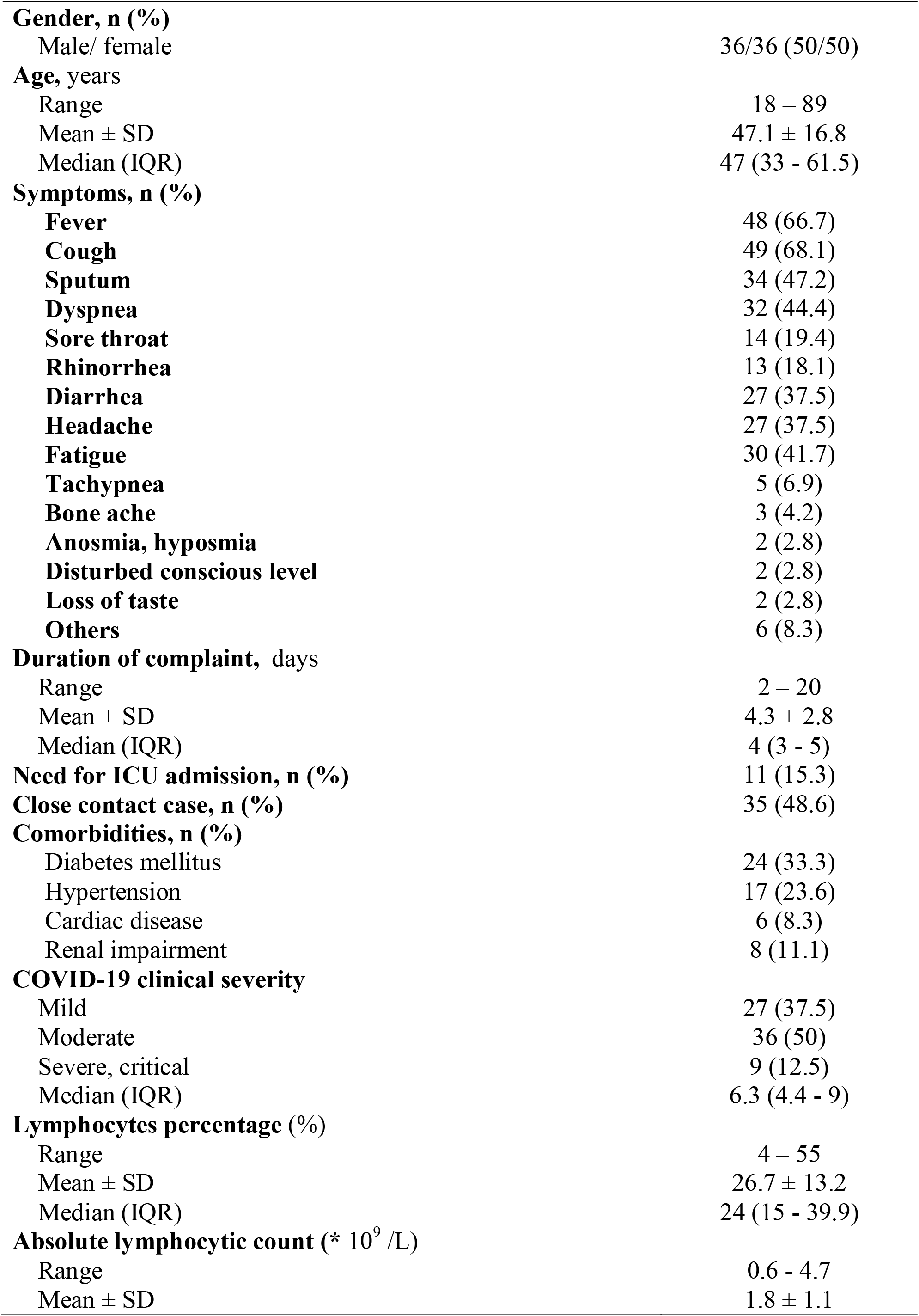
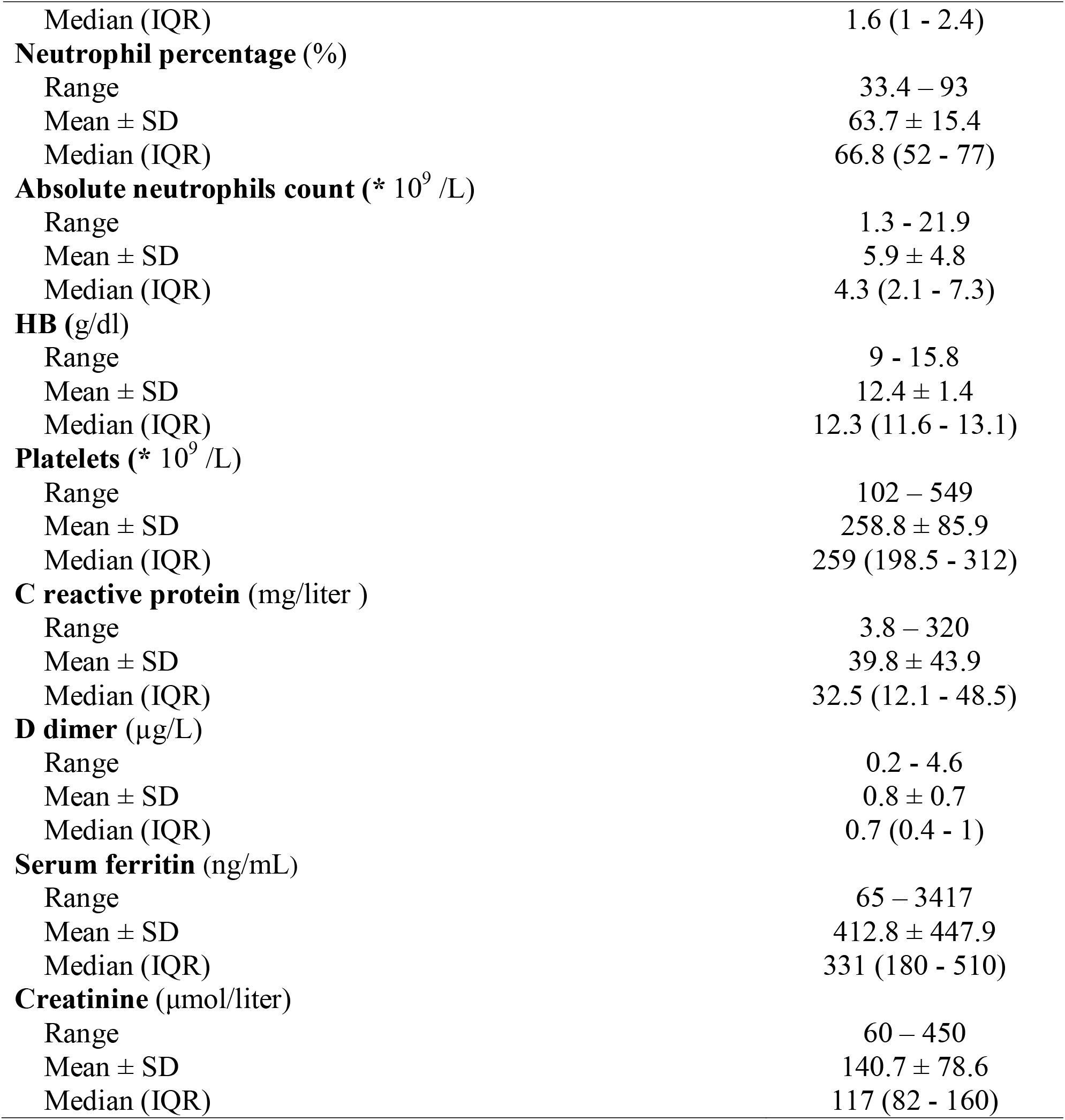
Demographic and clinical features of COVID-19 patients included in the study (n=72)

| Description (n=72) |
| --- |

|  |  |
| --- | --- |
| **Gender, n (%)** |  |
| Male/ female | 36/36 (50/50) |
| **Age, years** |  |
| Range | 18 – 89 |
| Mean ± SD | 47.1 ± 16.8 |
| Median (IQR) | 47 (33 - 61.5) |
| **Symptoms, n (%)** |  |
| **Fever** | 48 (66.7) |
| **Cough** | 49 (68.1) |
| **Sputum** | 34 (47.2) |
| **Dyspnea** | 32 (44.4) |
| **Sore throat** | 14 (19.4) |
| **Rhinorrhea** | 13 (18.1) |
| **Diarrhea** | 27 (37.5) |
| **Headache** | 27 (37.5) |
| **Fatigue** | 30 (41.7) |
| **Tachypnea** | 5 (6.9) |
| **Bone ache** | 3 (4.2) |
| **Anosmia, hyposmia** | 2 (2.8) |
| **Disturbed conscious level** | 2 (2.8) |
| **Loss of taste** | 2 (2.8) |
| **Others** | 6 (8.3) |
| **Duration of complaint, days** |  |
| Range | 2 – 20 |
| Mean ± SD | 4.3 ± 2.8 |
| Median (IQR) | 4 (3 - 5) |
| **Need for ICU admission, n (%)** | 11 (15.3) |
| **Close contact case, n (%)** | 35 (48.6) |
| **Comorbidities, n (%)** |  |
| Diabetes mellitus | 24 (33.3) |
| Hypertension | 17 (23.6) |
| Cardiac disease | 6 (8.3) |
| Renal impairment | 8 (11.1) |
| **COVID-19 clinical severity** |  |
| Mild | 27 (37.5) |
| Moderate | 36 (50) |
| Severe, critical | 9 (12.5) |
| Median (IQR) | 6.3 (4.4 - 9) |
| **Lymphocytes percentage (%)** |  |
| Range | 4 – 55 |
| Mean ± SD | 26.7 ± 13.2 |
| Median (IQR) | 24 (15 - 39.9) |
| **Absolute lymphocytic count (\* 10<sup>9</sup> /L)** |  |
| Range | 0.6 - 4.7 |
| Mean ± SD | 1.8 ± 1.1 |
| Median (IQR) | 1.6 (1 - 2.4) |
| **Neutrophil percentage (%)** |  |
| Range | 33.4 – 93 |
| Mean ± SD | 63.7 ± 15.4 |
| Median (IQR) | 66.8 (52 - 77) |
| **Absolute neutrophils count (\* 10<sup>9</sup> /L)** |  |
| Range | 1.3 - 21.9 |
| Mean ± SD | 5.9 ± 4.8 |
| Median (IQR) | 4.3 (2.1 - 7.3) |
| **HB (g/dl)** |  |
| Range | 9 - 15.8 |
| Mean ± SD | 12.4 ± 1.4 |
| Median (IQR) | 12.3 (11.6 - 13.1) |
| **Platelets (\* 10<sup>9</sup> /L)** |  |
| Range | 102 – 549 |
| Mean ± SD | 258.8 ± 85.9 |
| Median (IQR) | 259 (198.5 - 312) |
| **C reactive protein (mg/liter )** |  |
| Range | 3.8 – 320 |
| Mean ± SD | 39.8 ± 43.9 |
| Median (IQR) | 32.5 (12.1 - 48.5) |
| **D dimer (μg/L)** |  |
| Range | 0.2 - 4.6 |
| Mean ± SD | 0.8 ± 0.7 |
| Median (IQR) | 0.7 (0.4 - 1) |
| **Serum ferritin (ng/mL)** |  |
| Range | 65 – 3417 |
| Mean ± SD | 412.8 ± 447.9 |
| Median (IQR) | 331 (180 - 510) |
| **Creatinine (μmol/liter)** |  |
| Range | 60 – 450 |
| Mean ± SD | 140.7 ± 78.6 |
| Median (IQR) | 117 (82 - 160) |

### CT chest conclusions

***Table 2*** found that 67 (93.1%) patients of the total study population had CT chest anomalies. The baseline lesions identified were ground glass opacification identified in (67 cases, 93%), consolidation alone was observed in (22 cases, 22.2%), while consolidation with ground glass opacification was identified in (22 cases, 22.2%); most of the lesions were bilateral and multi-focal. The most commonly affected part was the left lower division. Moreover, thickening of the inter-lobular septa was observed in 16 cases (22.2%), vascular enlargement observed in 28 (38.9%), while one patient had pleural effusion, and the typical finding were “crazy paving sign” (19 cases, 26.4%) and “spider web sign” (12 cases, 16.6%), while halo sign was detected in 6 cases (8.3). The average PII value was (35.4 ± 14.7%) for all the cases.

**Table 2.** Radiological topographies of the COVID-19 population included in the study (n=72)

| **Chest CT findings of the population under study** | **Frequency, n (%)** |
| --- | --- |
| **Normal CT chest** | 5 (6.9) |
| **Presence of ground glass opacification** | 67 (93.1) |
| **Distribution of lesions** |  |
| Peripheral lesions | 47 (65.2) |
| Patchy lesions | 20 (27.7) |
| Bilateral lesions | 58 (80.6) |
| Multi-focal lesions | 58 (80.6) |
| Uni-focal lesions | 9 (12.5) |
| **Number of lobes** |  |
| 0 | 5(6.9) |
| 1 | 4 (5.5) |
| 2 | 8 (11.1) |
| 3 | 12 (16.7) |
| 4 | 7 (9.7) |
| 5 | 36 (50) |
| ≤ 3 | 27 (37.5) |
| > 3 | 43 (59.7) |
| **Left upper lobe** | 47 (65.3) |
| **Left lower lobe** | 59 (81.9) |
| **Right upper lobe** | 52 (72.2) |
| **Right middle lobe** | 44 (61.1) |
| **Right lower lobe** | 60 (83.3) |
| **Bilateral upper lobes** | 40 (55.6) |
| **Bilateral lower lobes** | 56 (77.8) |
| **Ground glass opacification + consolidation** | 16 (22.2) |
| **Consolidation without ground glass opacification** | 16 (22.2) |
| **Crazy paving pattern** | 19 (26.4) |
| **Vascular enlargement** | 28 (38.9) |
| **Thickening of the inter-lobular septa** | 23 (31.9) |
| **Bronchial wall thickening** | 5(6.9) |
| **Air bronchogram** | 16 (22.2) |
| **Air trapping sign** | 2 (2.8) |
| **Reverse halo sign** | 3 (4.2) |
| **Presence of halo sign** | 6 (8.3) |
| **Spider web sign** | 12(16.6) |
| **Pleural effusion** | 1(1.3) |
| **Pulmonary Inflammation Index in chest CT (%)** |  |
| Range | 5 - 57.5 |
| Mean ± SD | 35.4 ± 14.7 |
| Median (IQR) | 37.5 (25 - 50) |

### The determinants for ICU admission need and correlation with PII

We found that the cases that required ICU admittance presented mainly with dyspnea (81.8 vs. 37.7% of those non requiring ICU, P=0.009), tachypnea (P=0.023) and had higher rate of comorbidities (renal impairment, P=0.016). This group had considerably lowered lymphocytes percentage (P=0.002) and hemoglobin levels (P=0.001), higher serum creatinine (P=0.020) and ferritin (P=0.034). There was also observed increase in levels of C reactive protein and D Dimer (though non-significant). Regarding the radiological conclusions, >3 lobes affection and higher PII were ominously recorded in those who required ICU admittance (P= 0.041 and 0.013) ***(Table 3***).

**Table 3.** Comparisons between groups according to the necessity for ICU admittance.

|  | ICU admission |  |  |
| --- | --- | --- | --- |
|  | Yes (n=11) | No (n=61) |  |
| **Gender, n (%)** |  |  |  |
| Male/female | 8/3 (72.7/27.3) | 28/33 (45.9/54.1) | 0.101 |
| **Age, years** |  |  |  |
| Range | 36 - 89 | 18 – 77 |  |
| Mean ± SD | 54.4 ± 14.4 | 45.8 ± 17 |  |
| Median (IQR) | 55 (45 - 58) | 46 (33 - 62) | 0.141 |
| **Clinical symptoms, n (%)** |  |  |  |
| Fever | 9 (81.8) | 39 (63.9) | 0.316 |
| Cough | 10 (90.9) | 39 (63.9) | 0.093 |
| Sputum | 7 (63.6) | 27 (44.3) | 0.236 |
| Dyspnea | 9 (81.8) | 23 (37.7) | **0.009** |
| Sore throat | 4 (36.4) | 10 (16.4) | 0.207 |
| Rhinorrhea | 1 (9.1) | 12 (19.7) | 0.676 |
| Diarrhea | 7 (63.6) | 20 (32.8) | 0.088 |
| Headache | 2 (18.2) | 25 (41) | 0.191 |
| Fatigue | 2 (18.2) | 28 (45.9) | 0.107 |
| Tachypnea | 3 (27.3) | 2 (3.3) | **0.023** |
| Bone ache | 1 (9.1) | 2 (3.3) | 0.397 |
| Anosmia, hyposmia | 0 (0) | 2 (3.3) | 1.000 |
| Disturbed level of consciousness | 1 (9.1) | 1 (1.6) | 0.284 |
| Loss of taste | 0 (0) | 2 (3.3) | 1.000 |
| Others | 1 (9.1) | 5 (8.2) | 1.000 |
| **Duration of complains (Days)** |  |  |  |
| Range | 3 - 13 | 2 - 20 |  |
| Mean ± SD | 5.5 ± 3.1 | 4.1 ± 2.7 | |
| Median (IQR) | 4 (4 - 6) | 4 (3 - 4) | **0.028** |
| **Close contact case, n (%)** | 3 (27.3) | 32 (52.5) | 0.124 |
| **Comorbidities, n (%)** |  |  |  |
| Diabetes Mellitus | 5 (45.5) | 19 (31.1) | 0.488 |
| Hypertension | 3 (27.3) | 14 (23) | 0.714 |
| Cardiac disease | 1 (9.1) | 5 (8.2) | 1.000 |
| Renal impairment | 4 (36.4) | 4 (6.6) | 0.016 |
| **WBC (\* 10<sup>9</sup> /L)** |  |  |  |
| Range | 3.2 - 14 | 3.2 - 24.8 |  |
| Mean ± SD | 7.2 ± 3.1 | 7.5 ± 4.3 | |
| Median (IQR) | 6.5 (5.3 - 9) | 6.1 (4.3 - 9.1) | 0.876 |
| **Lymphocytes percentage (%)** |  |  |  |
| Range | 10 - 38 | 4 - 55 |  |
| Mean ± SD | 16.3 ± 8.3 | 28.6 ± 13 | |
| Median (IQR) | 15 (11 - 18) | 26.3 (18.7 - 40) | **0.002** |
| **Absolute lymphocytic count (\* 10<sup>9</sup> /L)** |  |  |  |
| Range | 0.6 - 1 | 0.6 - 4.7 |  |
| Mean ± SD | 0.8 ± 0.3 | 1.9 ± 1.1 | |
| Median (IQR) | 0.8 (0.6 - 1) | 1.7 (1 - 2.6) | 0.214 |
| **Neutrophil percentage (%)** |  |  |  |
| Range | 69 - 81 | 33.4 - 93 |  |
| Mean ± SD | 75 ± 8.5 | 63.2 ± 15.5 | |
| Median (IQR) | 75 (69 - 81) | 65 (52 - 76) | 0.316 |
| **Absolute neutrophils count (\* 10<sup>9</sup> /L)** |  |  |  |
| Range | 2.8 - 4.6 | 1.3 - 21.9 |  |
| Mean ± SD | 3.7 ± 1.3 | 6.1 ± 4.9 | |
| Median (IQR) | 3.7 (2.8 - 4.6) | 4.3 (2.1 - 7.8) | 0.742 |
| **Hemoglobin (g/dl)** |  |  |  |
| Range | 9 - 12 | 9.9 - 15.8 |  |
| Mean ± SD | 11 ± 1.2 | 12.6 ± 1.3 | |
| Median (IQR) | 11.7 (10 - 12) | 12.8 (11.8 - 13.2) | **0.001** |
| **Platelets (\* 10<sup>9</sup> /L)** |  |  |  |
| Range | 102 - 334 | 107 - 549 |  |
| Mean ± SD | 224.8 ± 83.7 | 264.9 ± 85.6 | |
| Median (IQR) | 236 (120 - 310) | 263 (199 - 312) | 0.280 |
| **C reactive protein (mg/liter )** |  |  |  |
| Range | 12.3 - 320 | 3.8 - 116 |  |
| Mean ± SD | 71.8 ± 87.9 | 34 ± 27.8 | |
| Median (IQR) | 45 (23 - 102) | 32 (12 - 45) | 0.066 |
| **D dimer (mg/liter )** |  |  |  |
| Range | 0.3 - 4.6 | 0.2 - 2.8 |  |
| Mean ± SD | 1.3 ± 1.4 | 0.7 ± 0.5 | |
| Median (IQR) | 0.9 (0.4 - 1.3) | 0.7 (0.4 - 0.9) | 0.161 |
| **Serum ferritin (ng/ml)** |  |  |  |
| Range | 199 - 1115 | 65 - 3417 |  |
| Mean ± SD | 552.3 ± 299.9 | 391 ± 465.4 | |
| Median (IQR) | 513 (312 - 727) | 321 (176 - 448) | **0.034** |
| **Creatinine (μmol/liter)** |  |  |  |
| Range | 60 - 450 | 65 - 312 |  |
| Mean ± SD | 247.3 ± 135.4 | 124.3 ± 50.8 | |
| Median (IQR) | 260.5 (128.5 - 345) | 113.5 (77.5 - 159.5) | **0.020** |
| **Radiological Findings, n (%)** |  |  |  |
| Bilateral lesions | 10 (90.9) | 48 (78.7) | 0.679 |
| Multi-focal lesions | 9 (81.8) | 49 (80.3) | 1.000 |
| Number of lobes affected |  |  |  |
| ≤ 3 | 1 (9.1) | 28 (45.9) | **0.041** |
| > 3 | 10 (90.9) | 33 (54.1) |  |
| **PII chest CT value (%)** |  |  |  |
| Range | 27.5 - 57.5 | 5 - 55 |  |
| Mean ± SD | 45.5 ± 8.9 | 33.4 ± 14.9 | |
| Median (IQR) | 47.5 (37.5 - 52.5) | 33.8 (24.5 - 46.3) | **0.013** |
| **Clinical Severity, n (%)** |  |  |  |
| Mild | 0 (0) | 27 (44.3) | **<0.001** |
| Moderate | 2 (18.2) | 34 (55.7) |  |
| Severe | 9 (81.8) | 0 (0) |  |

There were moderate positive correlation between PII chest CT value with age (r=0.264, P=0.031) and other prognostic inflammatory markers as ferritin (r=0.225, P=0.048), D Dimer (r=0.271, P=0.043) and serum creatinine as shown in ***Table 4***.

**Table 4.**
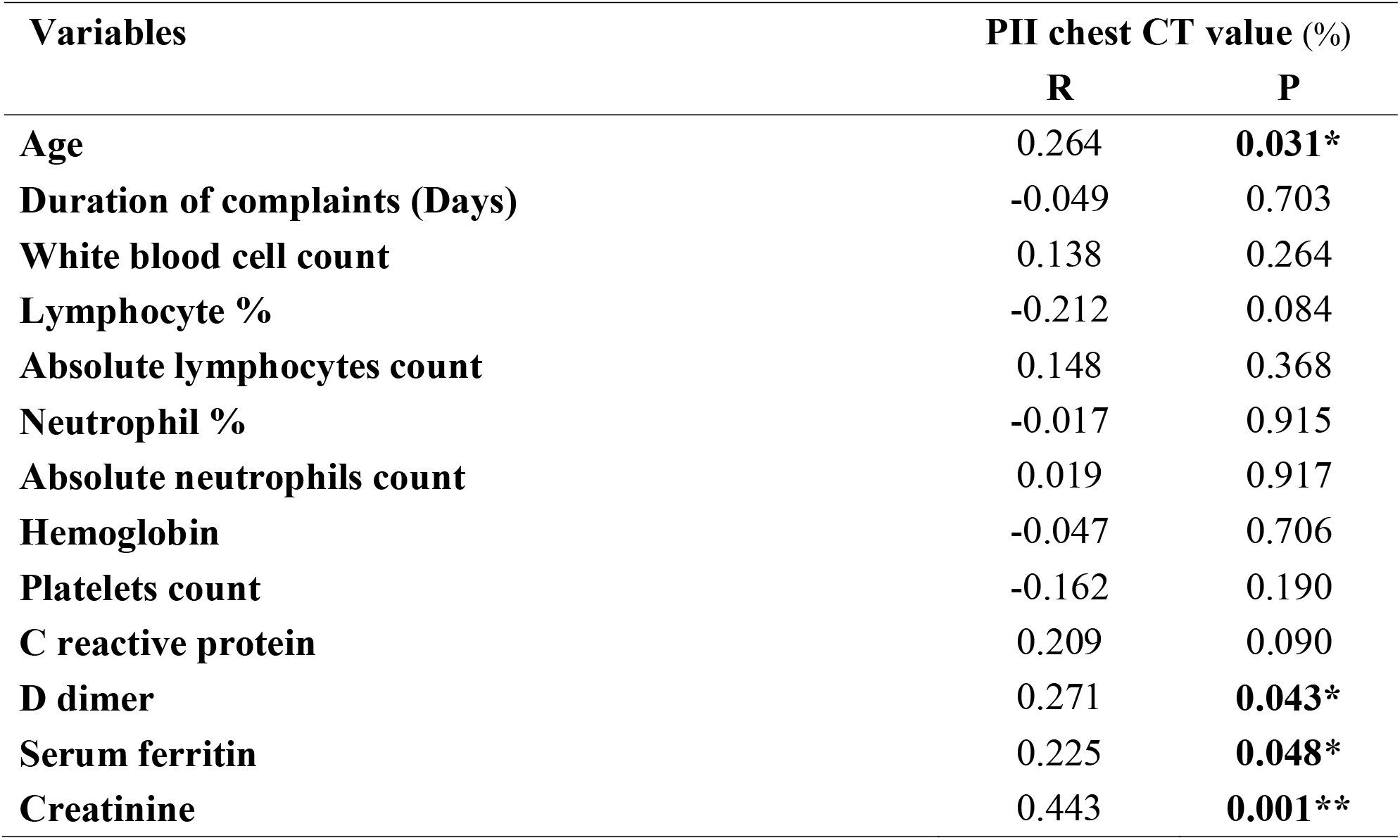
Correlation between Pulmonary inflammation index (PII) with clinical and laboratory prognostic parameters.

***Figure 1*** shows a HRCT chest of male patient, asthmatic, presented with fever, productive cough, dyspnea, tachycardia of 3 days. He was diagnosed as COVID-19, admitted to ICU. The CT findings are multi-lobular bilateral GGO with consolidation, crazy paving pattern, PII= 60%. The patient rapidly deteriorated and died.

**Figure 1.**
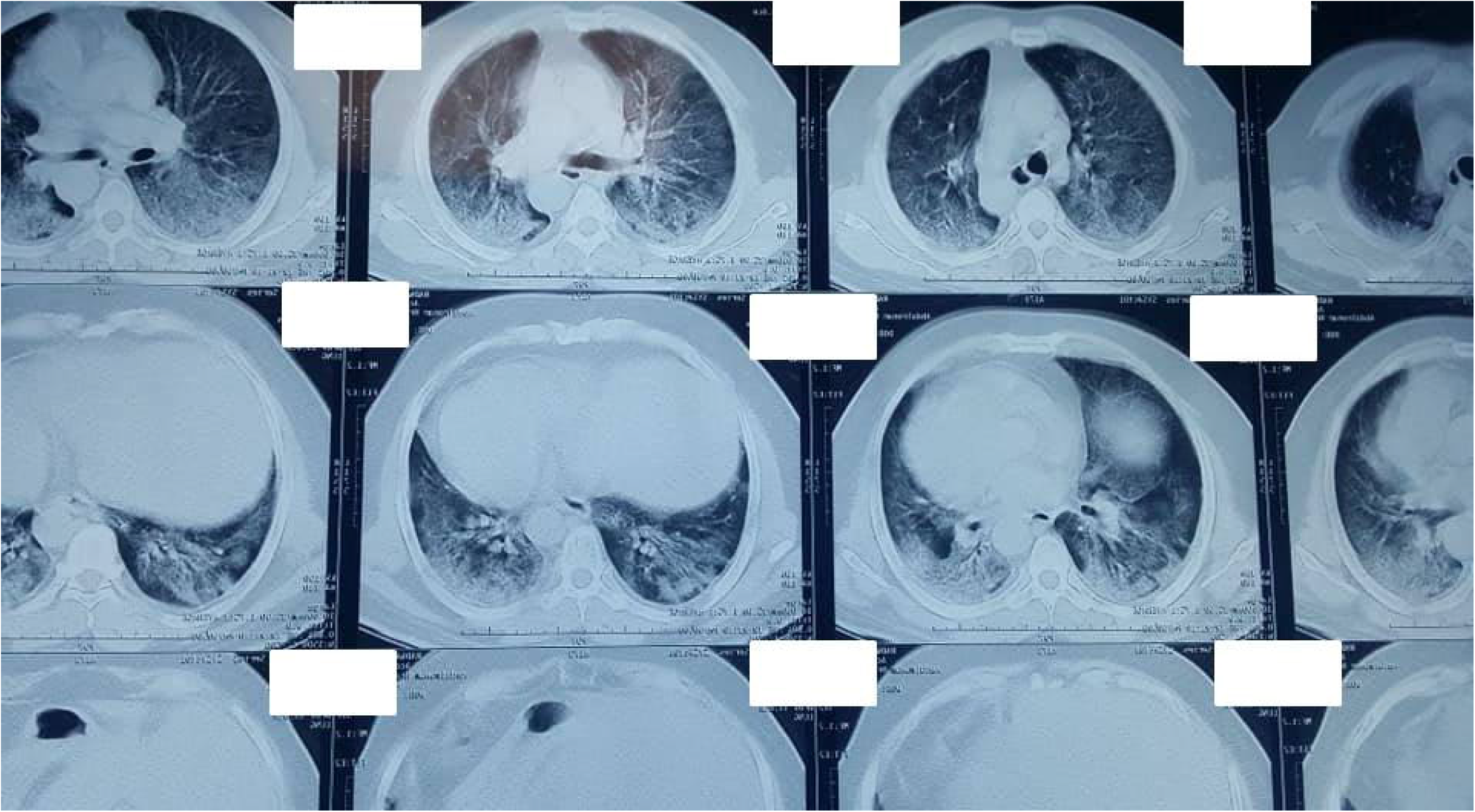
HRCT chest of male patient, asthmatic, presented with fever, productive cough, dyspnea, tachycardia of 3 days. He was diagnosed as COVID-19, admitted to ICU. The CT findings are multi-lobular bilateral GGO with consolidation, crazy paving pattern, PII= 60%. The patient rapidly deteriorated and died.

## Discussion

We performed an inclusive study of the demographic, clinical, laboratory and radiological topographies of 72 consecutive Egyptian COVID-19 patients admitted to our hospital. The main findings were the predominance of HRCT findings in majority of COVID-19 patients in different disease severity. The use of radiologic PII seemed very informative index of state of inflammation and correlated with prognostic parameters such as levels of ferritin and D dimer.

The median age in the study population was 47 years and it is well-matched with previous studies in China and United States (**9, 10**). There was also no obvious gender predilection in the current study. However, male predominance (up to 73%) was recorded repeatedly in several studies and was explained by more exposure to virus in outdoor working men (**11**). Furthermore, it was reported that one third of the positive COVID-19 cases had comorbidities including diabetes mellitus, hypertension and cardiac problems similar to our results (3, 12).

Regarding the clinical severity of the disease, most of the cases included in the study presented with moderate COVID-19 disease severity similar to Zu and colleagues (13). As expected the most common symptom among the studied population was cough (68.1), which is in harmony, with numerous reports (**6, 14, 15**). Diarrhea was evident in 37% of the cases comparable to reports recording that over 1/3 of the patients had symptoms suggesting gastro-intestinal affection such as diarrhea, nausea and vomiting (16).

Like early published reports, 5 cases (6.9%) in this study presented with normal CT chest even with positive PCR result suggesting that PCR result can be positive even in cases presented with normal CT findings (**5**). The normal chest CT findings frequency ranged from 0.7% to 56%, varied according to the disease severity with the highest rates reported in those with asymptomatic or with mild presentations (17).

Regarding the HRCT chest topographic features, we found that multi-lobar affection was noted in 80.6%, bilateral lower lobes affection in 83.3% and peripheral/sub-pleural affection in 65.2%. The common configurations were: ground glass opacification, consolidation, crazy paving pattern, vascular enlargement & interlobular septal thickening and the uncommon CT patterns were: reverse halo sign, pleural effusion, spider web sign, air trapping sign & bronchial wall thickening. These results universally approved with many published data **(13, 18, 19, 20)**. More than half of cases had multi-lobar affection and progression, and right lung was more commonly involved. Even when a single section was affected, the right lung was commonly involved. In the right lung, the lower section was commonly affected, while the upper section was often involved in the left lung **(21)**. This should specify that as the disease progress, the disease tend to distribute often all over both lung lobes.

Besides the role of HRCT in the detection of COVID-19, it also plays an essential role in the proper stratification and management of the disease. As other pneumonias, there was definite association between disease severity & chest CT topographies. Intensive care unit (ICU) cases on admittance often presented with bilateral multi-lobar and sub-segmental consolidations, compared with the non-severe cases who presented with bilateral GGOs and consolidation **(3)**. Chest CT might be Page 12 of 17 used as a monitor to detect the evolution of the coronavirus disease as patients with severe illness most commonly report bilateral and multi-lobar affection **(12,19,22,23, 24)**.

We found that the cases that required ICU admittance presented mainly with dyspnea, fatigue and had higher rate of comorbidities, lower lymphocytes percentage and hemoglobin levels while higher serum creatinine, D Dimer and ferritin. Decreased lymphocytes percentage on admittance was linked with worse consequences in cases with COVID-19 **(25)** and lymphopenia, in addition to the serum levels of D dimer, CRP, ferritin were considered as important factors in the hazard stratification of the severe and fatal form of COVID-19 in hospitalized cases **(26)** and high serum ferritin alone might be considered as a forecaster of poor outcome **(27, 28)**.

Cases with comorbid disorders signified more severe clinical signs compared to paralleled to those without any comorbid disorder (3). There is a strong relationship between the need for admittance to the intensive care unit and elevated score of qSOFA, higher levels of C-reactive protein and pro-calcitonin levels **(28, 29)**.

C-reactive protein (CRP) levels were increased in cases with COVID-19 and it has been revealed that survivors had greater levels compared with non - survivors, indicating a strong correspondence with the severity & prognosis of the disease **(30)**. This was in similar to the results of this study but the difference did not reach the level of significance (median 45 vs. 32, p=0.066). This may be attributed to the small number of ICU admitted patients in this cohort (only 11 cases).

A slight increase of D Dimer level was also noticed in this study in ICU cases (1.3±1.4 vs. 0.7±0.5). Greater levels of D-dimer may help in efficient differentiation between mild and severe cases of COVID-19 disorder **(16)** and may be used as a mirror for identification of disease progression toward unfavorable outcome and even death **(31,32)**.

We used the PII to quantify the lung lesions in patients with COVID-19 and found that the PII was ominously higher in cases requiring ICU admittance compared to those who did not. The PII was positively correlated with age, D Dimer, serum ferritin and creatinine. These findings may suggest the value of the PII as an independent index of early inflammation before clinical deterioration and it may predict the evolution from mild to severe infection **(33)**. Similar substantial correlations were observed between the degree of pulmonary inflammation and the main clinical symptoms and laboratory markers in COVID-19 patients **(6)**.

This study had some limitations. Firstly is restrospective design and the short duration for the collection of our records, secondly, we did not perform a follow-up CT to explore the radiology evolution of our cases, which might aid in the judgment of the course of the disease.

### Conclusions

We suggest that the use of PII together with clinical and laboratory data may be valuable in determining the inflammatory status of COVID-19. It was correlated to inflammatory markers as D dimer, ferritin even before clinical deterioration. This may allow clinicians to avoid advance of the disease by proper early intervention and improve cure rates.

## Data Availability

on request

## Notes

### Competing Interest Statement

The authors have declared no competing interest.

### Clinical Trial

NCT04479293 (secondry aim)

### Funding Statement

none

### Author Declarations

18-2020/14 (Ministry of Health and Population)

## References

1. Wang D, Hu B, Hu C, et al., Clinical characteristics of 138 hospitalized patients with 2019 novel coronavirus-infected pneumonia in Wuhan, China. Jama. 2020 Mar 17; 323(11): 1061–9.

2. Chung M, Bernheim A, Mei X, et al. CT imaging features of 2019 novel coronavirus (2019-nCoV). Radiology 2020.

3. Huang P, Liu T, Huang L, et al. Use of chest CT in combination with negative RT-PCR assay for the 2019 novel coronavirus but high clinical suspicion. Radiology 2020.

4. Xie X, Zhong Z, Zhao W, et al. Chest CT for typical 2019-nCoV pneumonia: relationship to negative RT-PCR testing. Radiology 2020. DOI: 10.1148/radiol.2020200343.

5. Ai T, Yang Z, Hou H, et al., Correlation of chest CT and RT-PCR testing in coronavirus disease 2019 (COVID-19) in China: a report of 1014 cases. Radiology. 2020 Feb 26:200642.

6. Wu C, Chen X, Cai Y, et al. Risk factors associated with acute respiratory distress syndrome and death in patients with coronavirus disease 2019 Pneumonia in Wuhan, China. JAMA Intern Med 2020. Doi: 10.1001/jamainternmed.2020.0994.

7. Hansell DM, Bankier AA, MacMahon H, et al. Fleischner society: glossary of terms for thoracic imaging. Radiology. 2008; 246:697–722.

8. Schoen K, Horvat N, Guerreiro NFC, et al. Spectrum of clinical and radiographic findings in patients with diagnosis of H1N1 and correlation with clinical severity. BMC Infect Dis. 2019; 19:964.

9. Singh AK, Gupta R, Misra A. Comorbidities in COVID-19: outcomes in hypertensive cohort and controversies with renin angiotensin system blockers. Diabetes Metab Syndr Clin Res Rev 2020. Doi: 10.1016/j.dsx.2020.03.016.

10. Richardson S, Hirsch JS, Narasimhan M, et al. Presenting characteristics, comorbidities, and outcomes among 5700 patients hospitalized with COVID-19 in the New York City area. JAMA 2020. Doi: 10.1001/jama.2020.6775.

11. Na Z, Ding Z, Wen W, et al. Clinical features of patients infected with 2019 novel coronavirus in Wuhan, China. Lancet 2020; 395: 497–605

12. Shi H, Han X, Jiang N, et al., Radiological findings from 81 patients with COVID-19 pneumonia in Wuhan, China: a descriptive study. The Lancet Infectious Diseases. 2020 Feb 24.

13. Zu ZY, Jiang MD, Xu PP, et al. Coronavirus disease 2019 (COVID-19): a perspective from China. Radiology 2020; 295(3):715–21.

14. Chen Q, Zheng Z, Zhang C, et al. Clinical characteristics of 145 patients with corona virus disease 2019 (COVID-19) in Taizhou, Zhejiang, China. Infection 2020. Doi: 10.1007/s15010-020-01432-5.

15. Guan WJ, Ni ZY, Hu Y, et al., Clinical characteristics of coronavirus disease in China. New England journal of medicine. 2020 Apr 30; 382(18):1708–20.

16. Zhang JJ, Dong X, Cao YY, et al., Clinical characteristics of patients infected with SARS-CoV-2 in Wuhan, China. Allergy. 2020(Online ahead of print).doi: 10.1111/all.14238. PubMed PMID: 32077115.

17. Sun Z, Zhang N, Li Y, Xu X. A systematic review of chest imaging findings in COVID-19. Quantitative Imaging in Medicine and Surgery. 2020 May; 10(5): 1058.

18. Lomoroa P, Verdeb F, Zerbonia F et al. COVID-19 pneumonia manifestations at the admission on chest ultrasound, radiographs, and CT: single-center study and comprehensive radiologic literature review. Eur J Radiol Open 100231, 2020.

19. Salehi S, Abedi A, Balakrishnan S, Gholamrezanezhad A. Coronavirus disease 2019 (COVID-19): a systematic review of imaging findings in 919 patients. American Journal of Roentgenology. 2020 Mar 14:1–7.

20. Raptis CA, Hammer MM, Short RG, et al. Chest CT and coronavirus disease (COVID-19): a critical review of the literature to date. American Journal of Roentgenology. 2020 Mar 26:1–4.

21. Awulachew E, Diriba K, Anja A, et al., Computed Tomography (CT) Imaging Features of Patients with COVID-19: Systematic Review and Meta-Analysis. Radiology Research and Practice. 2020 Jul 23; 2020.

22. Song F, Shi N, Shan F, et al., Emerging 2019 novel coronavirus (2019-nCoV) pneumonia. Radiology. 2020 Apr; 295(1):210–7.

23. Duan YN, Qin J. Pre-and post-treatment chest CT findings: 2019 novel coronavirus (2019-nCoV) pneumonia. Radiology. 2020 Apr; 295(1):21.

24. Sabri Y, Nasef A, Ibrahim I, et al., CT chest for COVID-19, a multicenter study-experience with 220 Egyptian patients. Egyptian Journal of Radiology and Nuclear Medicine (2020) 51:144. https://doi.org/10.1186/s43055-020-00263-6

25. Huang I, Pranata R. Lymphopenia in severe coronavirus disease-2019 (COVID-19): systematic review and meta-analysis. Journal of Intensive Care. 2019 Dec; 8(1):1–0.

26. Velavan TP, Meyer CG. Mild versus severe COVID-19: laboratory markers. International Journal of Infectious Diseases. 2020 Apr 25.

27. Mehta P, McAuley DF, Brown M, et al. COVID-19: consider cytokine storm syndromes and immunosuppression. Lancet; 2020.

28. Mohamed Hussein AA, Galal I, Mohamed M et al., Survival and 30-days hospital outcome in hospitalized COVID-19 patients in Upper Egypt: Multicenter study, July 2020, Preprint.

29. Almazeedi S, Al-Youha S, Jamal MH, et al., Characteristics, risk factors and outcomes among the first consecutive 1096 patients diagnosed with COVID-19 in Kuwait. EClinicalMedicine. 2020 Jul 4:100448.

30. Ruan Q, Yang K, Wang W, et al., Clinical predictors of mortality due to COVID-19 based on an analysis of data of 150 patients from Wuhan, China. Intensive Care Med. 2020

31. Zhou F, Yu T, Du R, et al. Clinical course and risk factors for mortality of adult inpatients with COVID-19 in Wuhan, China: a retrospective cohort study. Lancet. 2020; 395(10229): 1054–62.doi: 10.1016/S0140-6736(20)30566-3. PubMed PMID: 32171076. PubMed Central PMCID: PMC7270627.

32. Bashash D, Abolghasemi H, Salari S, et al. Elevation of D-Dimer, But Not PT and aPTT, Reflects the Progression of COVID-19 toward an Unfavorable Outcome: A Meta-Analysis. IJBC 2020; 12(2): 47–53.

33. Wan S, Yi Q, Fan S et al., Relationships among lymphocyte subsets, cytokines, and the pulmonary inflammation index in coronavirus (COVID-19) infected patients. British Journal of Haematology, 2020, 189, 428–437.

